# Trip duration drives shift in travel network structure with implications for the predictability of spatial disease spread

**DOI:** 10.1101/2020.10.27.20215566

**Authors:** John R Giles, Derek AT Cummings, Bryan T Grenfell, Andrew J Tatem, Elisabeth zu Erbach-Schoenberg, CJE Metcalf, Amy Wesolowski

## Abstract

Human travel is one of the primary drivers of infectious disease spread. Models of travel are often used that assume the amount of travel to a specific destination decays as cost of travel increases and higher travel volumes to more populated destinations. Trip duration, the length of time spent in a destination, can also impact travel patterns. We investigated the spatial distribution of travel conditioned on trip duration and find distinct differences between short and long duration trips. In short-trip duration travel networks, trips are skewed towards urban destinations, compared with long-trip duration networks where travel is more evenly spread among locations. Using gravity models imbedded in simulations of disease transmission, we show that pathogens with shorter generation times exhibit initial patterns of spatial propagation that are more predictable among urban locations, whereas longer generation time pathogens have more diffusive patterns of spatial spread reflecting more unpredictable disease dynamics.

## Introduction

During an infectious disease outbreak, anticipating where a pathogen will spread is an important part of planning an effective response^1,2^. The initial stages of an outbreak are typically characterized by spatial dynamics through the population from an introduction event, such as the 2014 Ebola outbreak in West Africa^3^ and the 2019 COVID-19 pandemic^4^. These outbreaks provide clear examples of how quickly a pathogen can spread through a highly susceptible population. The speed of spatial propagation depends not only on population susceptibility and intrinsic biological properties of the pathogen, but also on connectivity patterns driven by human travel. Being able to predict the spatial dynamics can help inform where and when interventions should be deployed^1,5^. Thus, mathematical models have been used to predict these patterns for a range of pathogens including Ebola^6–8^, influenza^9–11^, SARS-CoV-1^12–14^, and the ongoing COVID-19 pandemic caused by SARS-CoV-2 ^15–17^. Predicting disease spread in this manner presents unique challenges because mathematical models of disease spread often exhibit complex dynamics^18–20^ that have a limited prediction horizon^21^ and rely on detailed information about human mobility patterns^22–24^.

Spatial connectivity in such models frequently relies on data and/or models of human mobility^2,25^ which are derived from travel data such as travel surveys^26,27^, call data records^28–30^, air traffic data^7,31,32^ or App data^33,34^. In the absence of such data, mechanistic models of human movement are often used^35–38^. These data and models typically represent travel volume (raw number or relative magnitude of trips among locations per unit time) which is used as a proxy for actual travel patterns and to connect spatial locations, which is often represented as a metapopulation framework^39^. However, the manner in which travel volume scales directly with the rate of transmission among subpopulations depends on the specifics of epidemiological-relevant movement among locations and the potentially infectious contacts that occur between individuals^40–42^, which ultimately determines the extent to which human mobility patterns can explain spatial disease dynamics.

One way in which travel volume may not scale directly to transmission among subpopulations is if the amount of time that travelers spend in a destination (trip duration) varies spatially. Previous work has explored scaling transmission based on trip duration, typically as some return rate, which scales down transmission per route based on assumptions about travel behavior^35,43–46^. Return rate methods have been applied so that all routes are scaled down uniformly^35^ or according to the origin^43^, or in the case of Poletto et al.^45,46^ where return rate was determined functionally according to the degree distribution of the destination. It follows that the duration of trips may depend on the origin and destination location which would suggest that integrating trip duration data would not uniformly change spatial transmission patterns^47,48^. For example, studies of urban travel have found that trip duration is associated with the attractiveness of a destination (e.g. work, residence, recreation)^49–52^ and that differences among locations form discrete spatial regions with unique patterns of trip duration^53–55^. There is also a decay in travel volume with the cost of travel, which can be measured by physical characteristics of the travel network, such as geographic distance^48,56,57^, travel time^58^ or socio-economic^57,59^ differences among locations. To examine how spatial variability of trip duration alters connectivity and transmission, we analyzed mobility inferred using mobile phone data from Namibia. We find that, although short duration trips are strongly influenced by the cost of travel (measured by travel distance and population size), the cost of travel is less important for longer trips. Using measures of network heterogeneity and changepoint analysis, we show that this difference in perceived cost of travel changes the spatial distribution of trips, where short trip duration networks exhibit heterogeneous patterns of connectivity that shift to homogeneous patterns as trip duration increases. We find that gravity models can be adjusted to capture these different patterns and use these models to assess how this impacts the spatial predictability of transmission dynamics for a range of simulated pathogens.

## Results

We used an anonymized data set of call data records (CDRs) collected in Namibia from October 2010 to April 2014 to estimate mobility patterns. Previous studies have used versions of these CDRs and provide detailed descriptions of the data^48,60,61^. Briefly, mobile phone usage data from 2.2 million unique subscribers was used to estimate travel patterns based on the estimated daily location of each subscriber, where the daily location is determined by recording the most frequently used mobile phone tower for each subscriber each day^28,62^. A trip was recorded when the location of a subscriber changed from location A to location B on subsequent days, otherwise the subscriber was recorded as staying in location A. For each trip, the date of travel and the number of days the subscriber remained in the destination location (trip duration) was recorded. The locations of the mobile phone towers were then aggregated to the 105 administrative level 2 regions (districts) in Namibia. For each day, these data provide an estimate for the number of trips made among the 105 districts and the duration of each trip, which include 259.2 million trips during the 40 months of data collection.

To explore the impact of trip duration on connectivity patterns, we assessed travel by trip duration. Short-duration trips (e.g. trips lasting between 1-3 days) were characterized by a high proportion of trips to nearby and densely populated areas. There were fewer long-trip duration trips (30-60 days) that were more evenly distributed across all locations without a clear skew to nearby or higher population destinations (see Figure 2). These patterns change incrementally when assessed over a more continuous set of 20 duration-restricted subnetworks reflecting finer time intervals that are analogous to the generation times of several infectious pathogens (see Table 1). For each subnetwork, we calculated the statistical distributions of two network centrality measures: node strength (weighted degree per node) and node closeness (total weighted distance from all nodes), see Methods for more detailed description of these metrics. Although both measures capture overall network clustering, both have slightly different interpretations for epidemic dynamics. High node strength values occur when a subset of locations have higher than average number of trips which increases disease transmission since more travel among locations increase the likelihood of spatial spread. High node closeness values indicate the travel network has a few highly connected locations and many peripheral locations that are less connected. This type of structure can slow the rate spatial disease spread^63^. We found that for short-trip duration networks, both node strength and closeness values were high with an overall distribution that was heavy tailed, which implies that there are particular nodes (high population density urban locations) that are more highly connected (Figure S4). In contrast, the long-trip networks, had distributions of node strength and closeness that were lower and more uniform, which implies that these networks are less connected, and travel is more evenly distributed across all locations (see Figure 3A-B and S2).

**Table 1.** Table of trip duration intervals used, with number observations and total trip counts.

| Duration interval<br>(days) |  | Subnetwork | Description | Number<br>routes<br>observed | Number total trips<br>(10,000s) |
| --- | --- | --- | --- | --- | --- |
| Start | Stop |  |  |  |  |
| 0 | Inf | 0 | All durations | 10843 | 25926 |
| 0 | 1 | 1 | 0 to 1 days | 10508 | 12083 |
| 1 | 2 | 2 | 1 to 2 days | 10629 | 16324 |
| 2 | 3 | 3 | 2 to 3 days | 10391 | 6376 |
| 3 | 5 | 4 | 3 to 5 days | 10379 | 4455 |
| 5 | 7 | 5 | 5 to 7 days | 10173 | 2169 |
| 7 | 10 | 6 | 7 to 10 days | 10127 | 1444 |
| 10 | 14 | 7 | 10 to 14 days | 9977 | 1076 |
| 14 | 21 | 8 | 2 to 3 weeks | 10013 | 863 |
| 21 | 28 | 9 | 3 to 4 weeks | 9740 | 475 |
| 30 | 60 | 10 | 1 to 2 months | 10044 | 617 |
| 60 | 90 | 11 | 2 to 3 months | 9274 | 203 |
| 90 | 120 | 12 | 3 to 4 months | 8505 | 98 |
| 120 | 150 | 13 | 4 to 5 months | 7594 | 49 |
| 150 | 180 | 14 | 5 to 6 months | 6892 | 29 |
| 180 | 210 | 15 | 6 to 7 months | 6173 | 19 |
| 210 | 240 | 16 | 7 to 8 months | 5706 | 13 |
| 240 | 270 | 17 | 8 to 9 months | 4943 | 9 |
| 270 | 300 | 18 | 9 to 10 months | 4379 | 6 |
| 300 | 330 | 19 | 10 to 11 months | 4052 | 5 |
| 330 | 360 | 20 | 11 to 12 months | 3819 | 4 |

**Figure 1.**
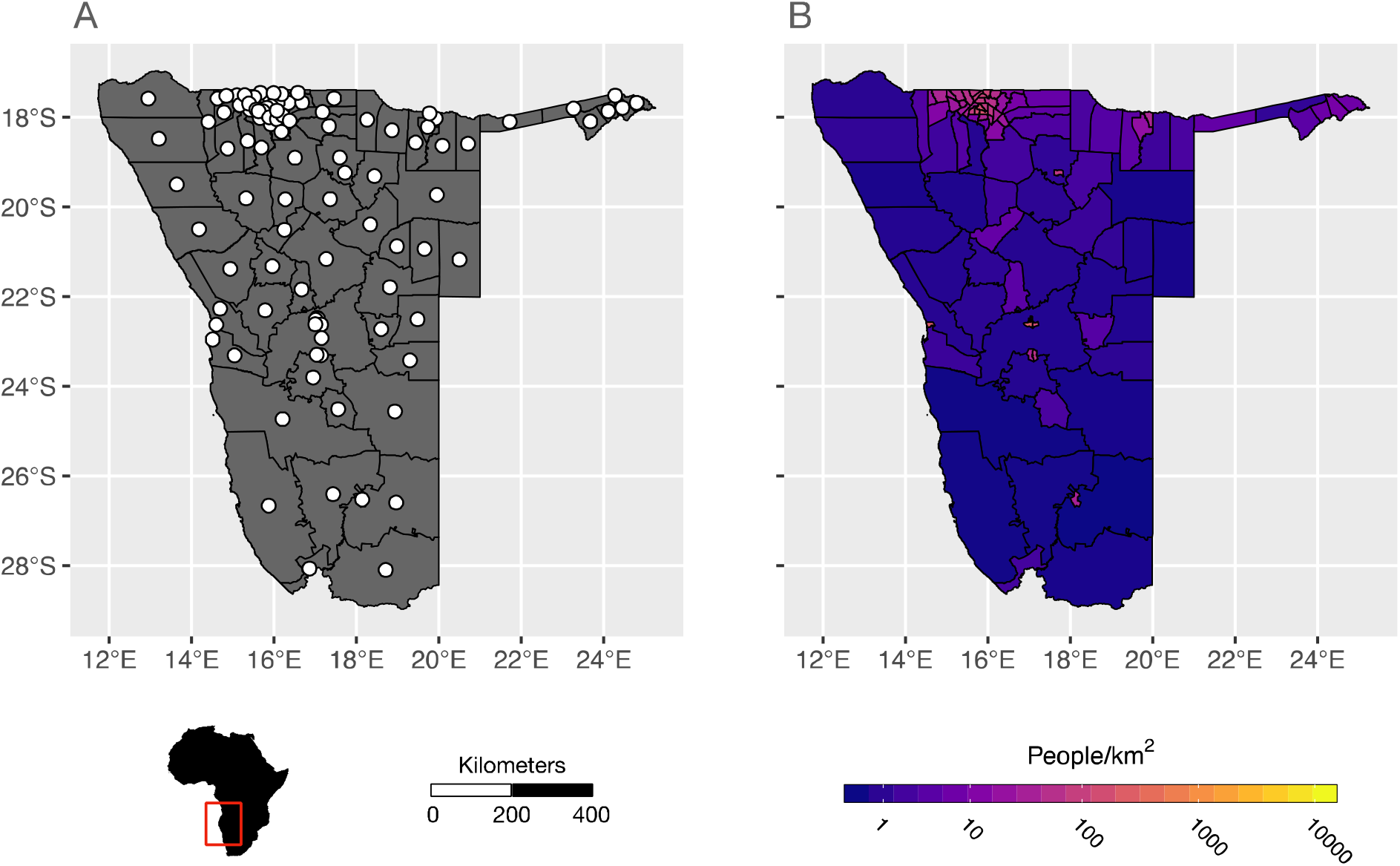
The Administrative districts and population distribution in Namibia. A) shows the locations of the 105 districts (administrative level 2) in Namibia with district centroids of each shown with white circles. B) The population density of each district in calculated as the number of people per square kilometer from the 2010 WorldPop Project estimates of the total number of people per 100m grid cell within each district (www.worldpop.org).

**Figure 2.**
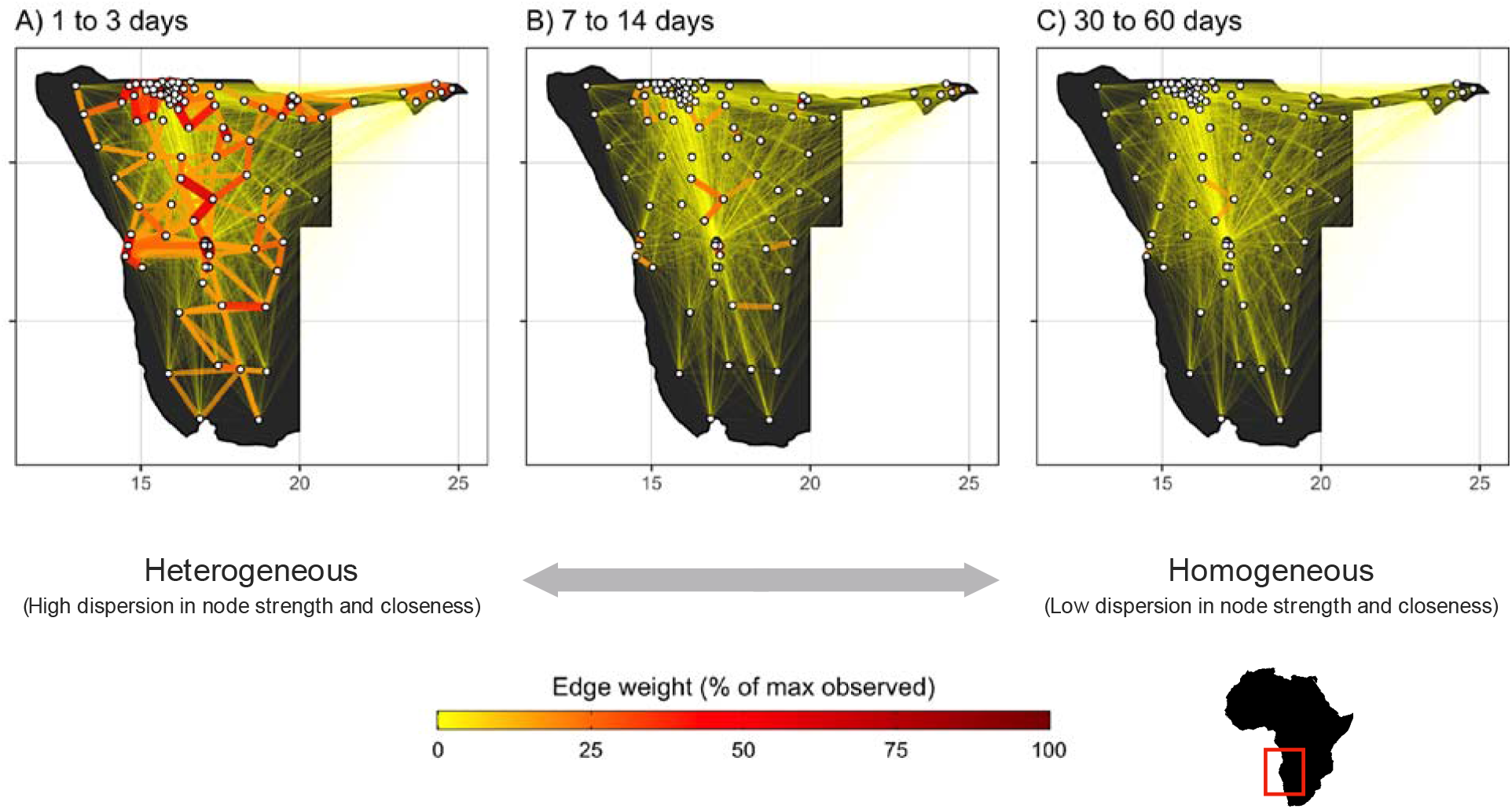
Travel network topology shifts from heterogeneous to homogeneous as trip duration increases. Maps of Namibia with travel volumes between districts that fall within three broad intervals of trip duration: A) 1-3 days, B) 7-14 days, and C) 30-60 days. For comparison, connectivity is defined as relative edge weight from 0 to 100%, which is calculated by scaling trip volume along the edges in each sub-network by the overall maximum trip volume observed in the full travel network. District centroids (nodes of the network) are indicated by the white circles.

**Figure 3.**
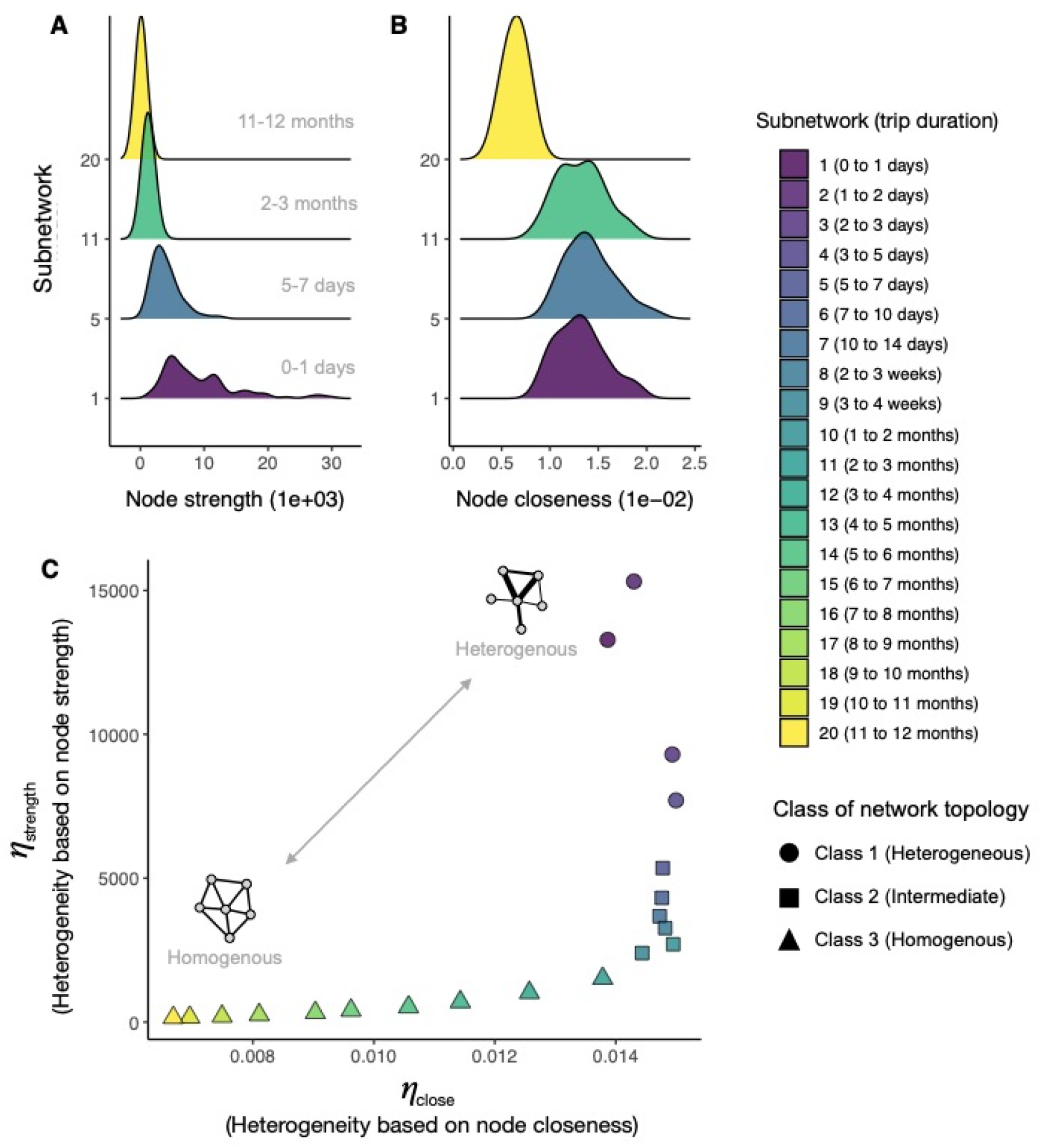
Distribution of node strength and node centrality for models with different trip duration intervals and changepoint analysis showing which trip durations constitute shifts in network topology. The empirical distributions of A) node strength (weighted degree per node) and B) node closeness (total weighted distance from all nodes) are plotted for four duration-restricted subnetworks which include trips of 1) 0-1 days, 5) 5-7 days, 11) 2-3 months, and 20) 11-12 months. C) The joint distribution of *η*_strength_ and *η*_close_ which measures the amount of network heterogeneity and structure based on the distributions of node strength and node closeness respectively. Each point represents one of the duration-restricted subnetworks and is colored according the duration interval shown in the color key to the right. The multivariate changepoint algorithm identified two significant shifts in network topology based on the joint distribution of *η*_strength_ and *η*_close_ that are placed at 5 and 60 days trip duration. The three nominal classes delineated by these thresholds are indicated by a circle for the heterogeneous class (1-5 days), a triangle for the homogeneous class (>60 days), and a square for intermediate class (6-60 days).

In general, analysis of these network centrality measures suggests that as trip duration increases, the spatial structure shifts from highly heterogenous connectivity (high level of variance in node strength and closeness) concentrated among urban centers to more homogenous connectivity (same overall degree distribution and lower closeness values). To more directly quantify the manner of this shift, we used a multivariate changepoint algorithm to identify statistically significant changes in node strength (η_strength_) and node closeness (η_close_) that indicate durations of travel at which network topology shifts significantly from heterogeneous to homogenous. We identified statistically significant shifts in network topology at 5 and 60 days (*p*-value = 1e-03 and 3e-04 respectively), which divides the duration-restricted subnetworks into three nominal classes: heterogeneous (1-5 days), intermediate (6-60 days), and homogeneous (>60 days; see Figure 3C). Overall, these results suggest that trips with a duration of less than 5 days display connectivity that is highly clustered among densely populated areas (Class 1 in Figure 3C), but trips longer than 60 days have more homogenously distributed trip volume among all locations (Class 3 in Figure 3C). In between these extremes is an intermediate class where trips are more evenly distributed among all locations, but there is still some clustering among more densely populated locations (Class 2 in Figure 3C). These results suggest that pathogens whose generation time is within the broad trip duration interval of the Class 1 heterogeneous topology (1-5 days) will depend more on short-trip networks for spatial propagation and may therefore exhibit patterns of disease spread along major routes of travel among urban population centers. Whereas pathogens with longer generation times may see more diffusive patterns of spread that are not dominated by urban travel.

To understand factors driving differences in connectivity, we examined trip counts within each duration-restricted subnetwork and its dependence on: i) distance between origin and destination, and ii) the destination’s population density. Using log-linear models, we estimated the effect size (fitted slope) and found that distance had a negative effect consistent with trip counts that decay as distance increases and destination population density had a net positive effect (i.e. more trips attracted to densely populated places). Since these covariates have rather different scales, we standardized them based on their standard deviation to allow for comparison and we found that on average, distance had an effect size approximately 10 times larger than destination population density (mean effect size across locations = −1.11 and 0.11 respectively), indicating that in largely rural Namibia, distance remains the primary driver of trip volume regardless of trip duration. We also found that effect sizes for both covariates were strongest for short duration trips (≈ 5 days or less), but these effects are reduced to near-zero for longer trip durations (Figure 4C and D), and for some origin districts the effect of destination population was reversed for longer trips (> 60 days), which suggests that districts with lower population density may even attract more long duration trips (Figure 4B and D).

**Figure 4.**
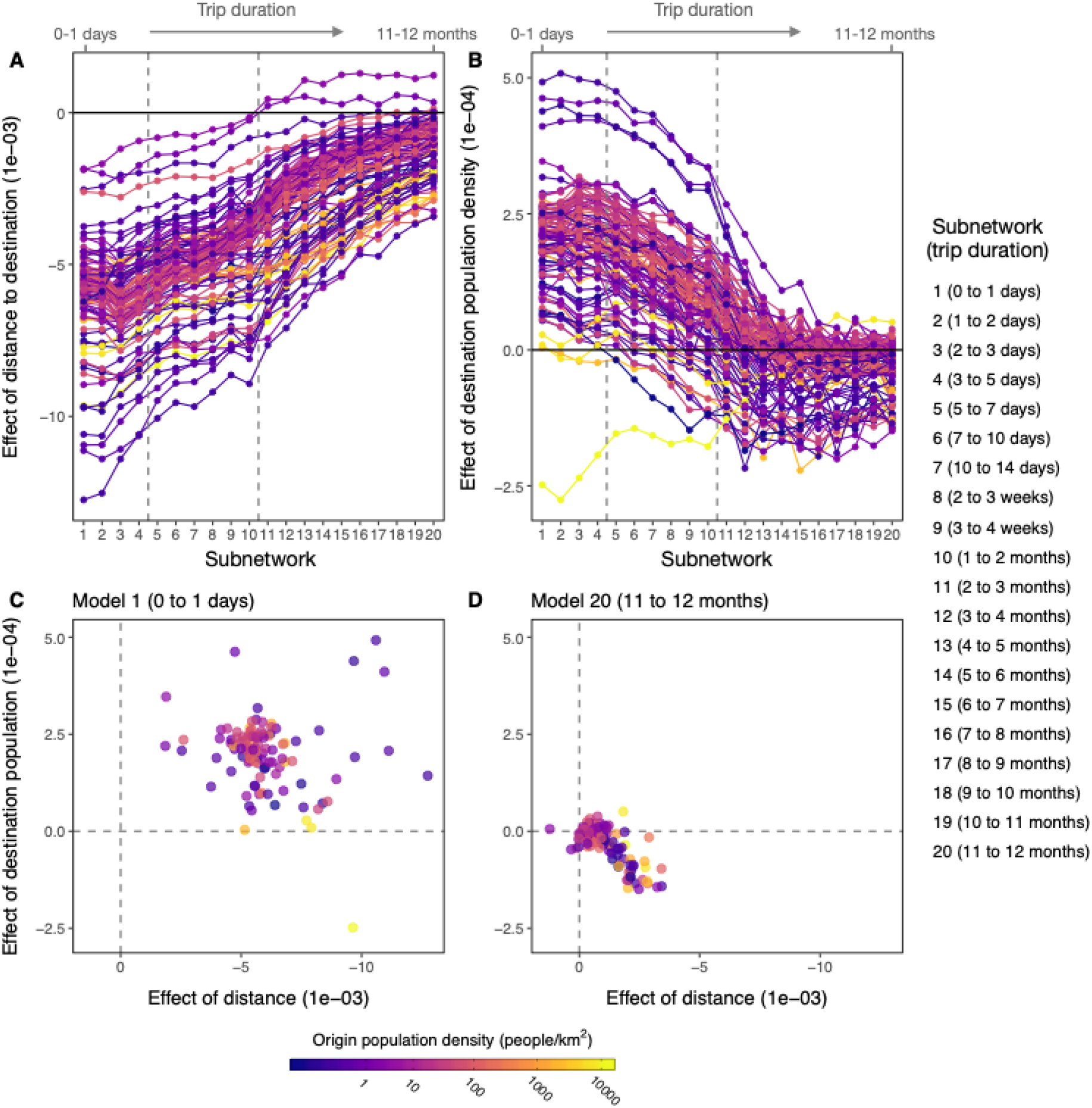
The drivers of trip volume across varying trip durations. The effect of distance to destination (A) and destination population size (B) on trip volume plotted across the 20 duration-restricted subnetworks. Each colored line represents one of the 105 districts with the population density of the origin district is indicated by the color bar. Dashed vertical lines indicate the network topology thresholds identified by the changepoint analysis. Scatterplots showing the joint distribution of effect sizes for distance (x-axis) and destination population (y-axis) for C) the model with the shortest duration (Model 1, 0-1 days) and D) the model with the longest duration (Model 20, 11-12 months). Comparison of C and D show that the effect size of both covariates is reduced to near-zero effect for longer trip durations.

A simple gravity model, the most commonly used spatial interaction model, was able to capture the changes in spatial connectivity over each of the 20 duration-restricted subnetworks (Table 1). When we compared gravity models fitted to each subnetwork to that of a full travel network with all trips, we found that the duration-restricted models are similar to the full model containing all trip durations up to 7-10 days duration (subnetwork 6), after which both the number of trips and connectivity values estimated by the gravity models begin to decrease, becoming essentially uniformly distributed after 5-6 months (subnetwork 14; see Figure 5A). The changes in spatial connectivity in these models result from incremental decreases in the distance parameter γ and destination population size parameter ω_2_, where both were essentially zero after 5-6 months duration (Figures 5B and C), indicating that distance and population size have little influence on the spatial distribution of these longer duration trips. Further, model fit decreased as trip duration increased, with R-squared values showing good model fit up to 1-2 months duration (subnetwork 10; R-squared = 0.51-0.55), but goodness of fit decreased for longer durations (see Figure S6). However, since the observed number of routes and trips decrease for trips of longer durations (Table 1 and Figure S1), this could spuriously cause the observed decrease in model fit (Figure S6). To ensure that connectivity patterns estimated by these models were not just due to the reduced number of observed routes or trip counts, we artificially down-sampled the full data according to the observed routes and sample sizes in Table 1. We found that the connectivity values estimated by gravity models fitted to artificially down-sampled subnetworks were largely the same as the full model (Figures S7 and S8), which indicates that the difference in spatial patterns among the actual subnetworks are robust to the smaller numbers of observations or trip counts.

**Figure 5.**
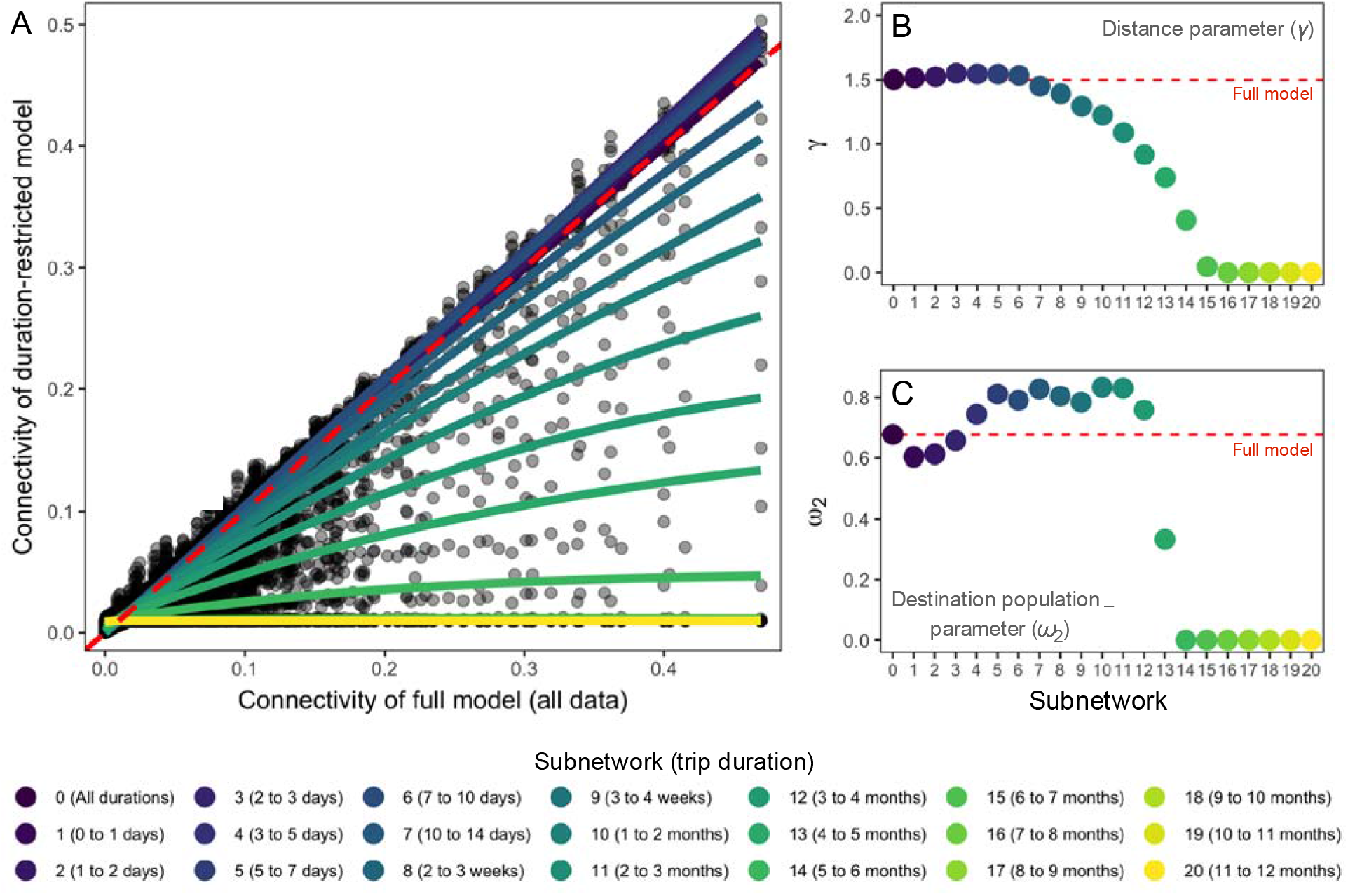
Change in connectivity and gravity model parameters fitted to travel data with increasing duration intervals compared to full model. A) The distribution of connectivity values for duration-restricted models (y-axis) in comparison to the full model that includes all data (x-axis). The smoothed lines indicate the change in connectivity for duration-restricted models with larger duration intervals showing a more evenly distributed pattern across all locations compared to the null model. The dashed red line indicates connectivity values that are equal to the full model. In B) and C), the change in fitted gravity model parameters (distance parameter γ and destination population parameter ω_2_ respectively) for increasing trip duration intervals. The color gradient indicates the duration interval of each model and the dashed red line shows the fitted parameter value for the full model, which includes all trip durations.

### Predictability of spatial spread

Changes in network topology are likely to impact the pattern of spatial spread for pathogens based on the time scale of transmission. For example, subnetworks for longer-duration trips have more uniform spatial distribution which means that many locations are more evenly connected. Intuitively, this would impact the predictability of where a disease might spread because there is limited spatial structure and there is not a single destination that stands out as having higher risk of importation based on travel alone. To explore this concept of predictability, we first modeled the spatial connectivity of disease spread for a range of pathogen life histories (Table S1) and then estimated the spatial force of infection conditioned on the generation time of each pathogen (see Methods). Starting with an initially infected location, we then assessed how reliably we can identify the next location where the disease would spread for each successive generation using an index of spatial predictability (ϕ). Spatial predictability ϕ is based on the principle of maximum entropy and provides an easily interpretable index between 0 and 1 that quantifies how the probability of disease importation is distributed among destinations—see Methods for detailed description.

When we analyzed the spatial simulations for each pathogen, we found that, given the same starting location (capital district of Windhoek), pathogens with shorter generation times and low *R*_0_ had the most predictable spatial dynamics in the initial stages of an outbreak (Figure 6A). In comparison, pathogens with longer generation times had less predictable spatial dynamics. However, this varied by pathogen characteristics where a high *R*_0_ can compensate for a short generation time making initial dynamics less predictable because these pathogens are able to spread to multiple destinations very quickly. Beyond initial dynamics, predictability decreased with successive generations for all pathogens although the rate of decrease varied by pathogen (Figure 6B). For example, measles and Ebola had comparable values of spatial predictability at the outset of the simulation due to their similar generation times. But in the case of measles, with a high *R*_0_, predictability declines more rapidly because it has a faster rate of transmission and associated spatial spread. This suggests that, while heterogeneity of the travel network has considerable effect on spatial predictability in initial outbreak stages, local dynamics driving the growth rate of the infected population in the index location (i.e. *R*_0_, proportion susceptible) determine the number of generations before spatial predictability rapidly decays.

**Figure 6.**
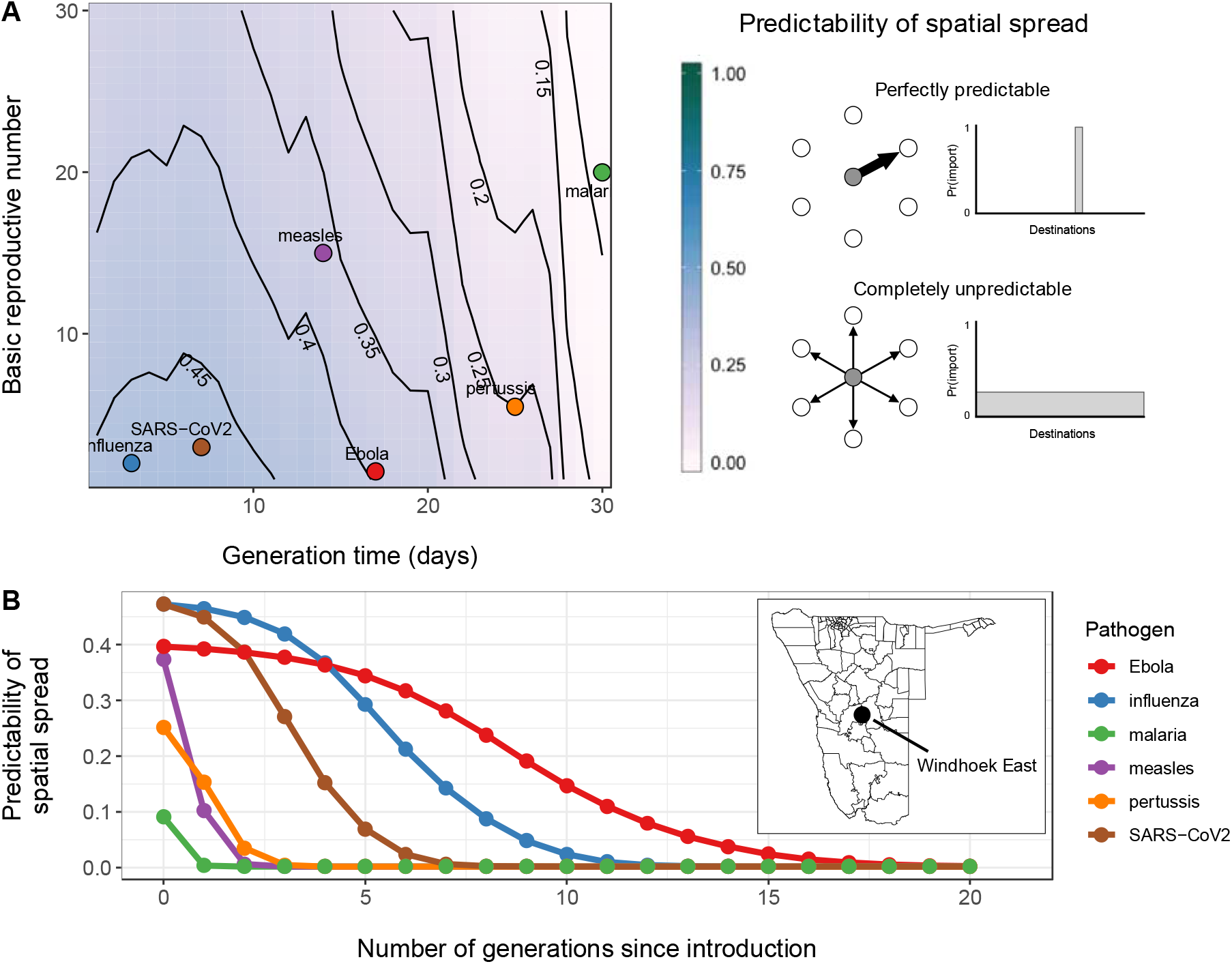
Predictability of spatial spread for a range of R_0_ and generation time values and the change in spatial predictability over time for 6 pathogens. A) A heatmap with contour lines showing the values of initial spatial predictability (ct) calculated for hypothetical combinations of R_0_ and generation time (days). Example pathogens (influenza, SARS-CoV-2, measles, Ebola, pertussis, and *P. falciparum* malaria) are indicated by the colored circles. The level of spatial predictability shown in the color bar to the right with schematic representations for scenarios where patterns of spatial spread are perfectly predictable (ϕ = 1) or completely unpredictable (ϕ = 0). B) The the change in these initial values of spatial predictability over successive generations for each of the 6 example pathogens. In both analyses, the capital district of Namibia, Windhoek East was used as the introduction district.

These patterns of spatial predictability were also dependent on the chosen starting location for the initial infected case in each simulation. For all pathogens, we found that predictability was sustained the longest for introduction into densely populated districts (Figure 7). Interestingly, we also found that the highest value of initial spatial predictability was *ϕ* = 0.63 (an influenza like simulation with an introduction even in the capital district) meaning that, given the connectivity patterns observed in Namibia, in the best-case scenario spatial spread appears to be marginally more predictable than unpredictable. We investigated this theoretically and found that a value of *ϕ* = 0.63 is functionally equivalent to the expectation that transmission is likely to occur from a given origin to 5 out of the 104 potential destinations (Figure S9A). In instances where the origin has more than one travel destination, which is common in highly connected travel networks, perfect certainty (*ϕ* = 1) is exceedingly unlikely (Figure S9B). Although these results suggest that predictability is higher for pathogens introduced into urban areas, in most cases we can expect that spatial spread in initial outbreak stages is generally more unpredictable than it is predictable and decreases with time.

**Figure 7.**
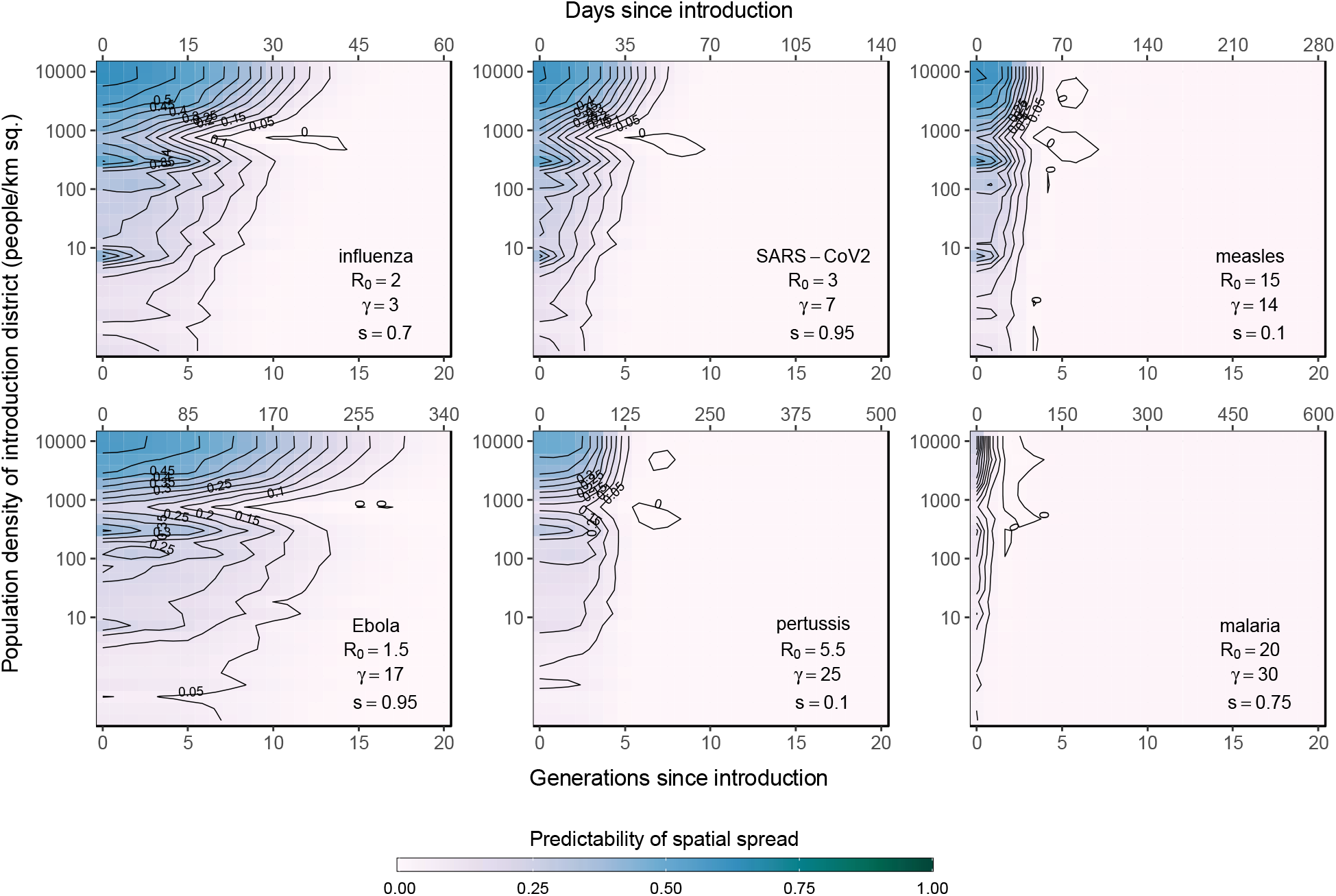
The change in spatial predictability for 6 pathogens plotted over time for outbreaks introduced to districts with a range of population densities. The results from simulations of outbreaks for 6 example pathogens (influenza, SARS-CoV-2, measles, Ebola, pertussis, and *P. falciparum* malaria). For each pathogen, outbreaks were introduced into each of the 105 districts in Namibia and the change in spatial predictability for successive generations was calculated. The heatmaps and contour lines show values of spatial predictability (ϕ) as they change over successive generations (x-axis) and the population density of the introduction district (y-axis). The number of days since the introduction is indicated on the top-axis. Annotations in the lower right indicate the pathogen, basic reproduction number (R_0_), generation time in days (γ), and proportion of the population that is susceptible (*s*) used in spatial simulations. Transmission parameters used in simulations were drawn from the literature and shown in Table S1.

## Discussion

There is often limited data about both the number and duration of trips to model spatial connectivity in disease transmission models. Previous work has shown that trip duration is subject to economic constraints due to the cost of travel^49,57^ and these constraints often depend on characteristics of the destination^49–52^. The impact of these types of travel decisions on spatial connectivity and disease transmission has been explored theoretically using metapopulation models, but few studies have quantified this effect empirically with data that span larger spatial and temporal scales^43–46,48^. However, we show that a travel network can be broken down into subnetworks based on trip duration. When we account for travel of different durations in these subnetworks, we find different spatial patterns in connectivity, where short-trip networks can be characterized by high node degree strength and node closeness with connectivity that is concentrated among densely populated areas. Conversely, long-trip networks can be characterized by lower node degree strength and node closeness, suggesting a spatial pattern of connectivity that is more evenly dispersed across all nodes, including those in low density locations. These spatial differences in network topology are likely driven by travel decisions stemming from the cost of travel, where the different classes of subnetworks emerge from variable economic constraints on trips depending on the duration^64^. Accordingly, we find that short trips are more constrained by the cost of travel, thus these trips concentrate around densely populated locations and show more heterogeneous structure like scale-free networks. But longer duration trips are less constrained by the cost of travel and therefore show more homogeneous network topology.

Spatial heterogeneity in trip duration impacts infectious disease spread differentially through interaction with generation times of each pathogen. Keeling and Rohani^44^ provide a theoretical precedent for the interdependence between pathogen life history and patterns of spatial connectivity. They show that when trip duration is shorter than the infectious period of the pathogen, interdependence between pathogen transmission parameters and coupling among locations increases. Accordingly, the spatial force of infection for pathogens with longer generation times is less skewed towards the distribution of shorter trips because the duration of shorter trips comprises a much smaller proportion of the total infectious period, allowing fewer opportunities for onward transmission. By comparison, longer duration trips contribute more towards the spatial force of infection for a long generation time pathogen because the duration of these trips has greater overlap with the infectious period, translating into more infectious days spent travelling all else being equal. This proportional difference effectively dilutes the force of infection for longer generation time pathogens in discrete-time models, because individuals are assumed to be infectious for the entire timestep. Spatial transmission in this manner is most strongly driven by trips with a duration approximately equal to or greater than the generation time of the pathogen^30^. Therefore, we see intrinsic spatial bias in the transmission process, where spatial spread of pathogens with shorter or longer generation times are potentially driven by vastly different patterns of spatial connectivity with fundamental effects on the overall spatial predictability of disease spread. For example, the spatial force of infection for a short generation time pathogen like influenza will be primarily driven by the short-trip duration subnetworks which have higher heterogeneity and clustering around high-density urban areas. By comparison, spatial transmission of pathogens with longer generation times (e.g. pertussis or malaria) will be primarily driven by the long-trip duration subnetworks that have more homogenous spatial structure and lower overall connectivity. Therefore, short generation time pathogens may exhibit more predictable patterns of spatial spread that disseminate predominantly through densely populated areas whereas longer generation time pathogens may exhibit more unpredictable patterns since the spatial force of infection is distributed more evenly across many destination locations.

Although we mainly investigated human mobility as the primary driver of spatial dynamics, the generalizability of our results to other disease systems also depends on spatial patterns in demographic and epidemiological factors specific to the pathogen. For example, transmission of childhood diseases, such as measles and rubella, depends on the rate at which children travel with adults and the demographic structure in each location. Heterogeneity in demographics among locations could change the patterns of spatial spread or introduce additional uncertainty. Population-level susceptibility also plays a significant role in the timing of spatial spread and is particularly important for pathogens with low population-level immunity^65,66^. Although we include a simple susceptibility parameter in our models of spatial spread, spatial heterogeneity in population susceptibility would also impact the locations that are at highest risk of spatial spread, as seen in instances where accumulation of susceptible individuals occurs due to vaccine refusal^67^ or spatially heterogeneous vaccination coverage^68^. Further, we used a discrete-time model with Susceptible-Infected-Recover disease dynamics which assumes individuals are uniformly infectious for the entirety of a timestep. However, it is less clear how the interdependence between trip duration and pathogen life history plays out when considering the precise timing of a trip in relation to different stages of disease (e.g. exposed, infectious, recovered) and whether infection has direct impact on the ability to travel. Addition work should be done to explore how the observed distribution of trip duration and uncertainty around latent period and infectious period may cause different dispersal dynamics depending on pathogen life history.

Methods for including trip duration have been proposed that include both theoretical^44,46^ and data-driven^48^ approaches. These methods offer a representation of spatial disease transmission that incorporates additional nuances of human travel decisions, but the resulting patterns of spatial spread may ultimately tend toward more unpredictable behavior (a persistent challenge when predicting the behavior of nonlinear systems in ecology and epidemiology^18–21^). Methods which account for uncertainty in human mobility and resulting disease dynamics therefore become especially important in order to make robust inferences about spatial transmission. For example, Kahn et al.^69^ showed that pathogens with longer incubation periods have patterns of spatial spread that are less predictable, which has important implications for vaccination campaigns. While the authors did not explicitly look at trip duration and network topology as we do here, these results lend support to our findings that the biological properties of pathogens that determine their speed of spatial propagation interact with the travel behavior of humans in a pathogen-specific manner, and suggests that this general relationship may be more widely applicable. However, the extent to which our results here fully generalize to disparate geographic settings is unclear due to the exceptionally low population density across much of Namibia with highly clustered populations in a few urban areas. This unique population distribution causes high variability in the size of administrative units, which are reflected in the physical placement of the network nodes in our analyses. The shifts in network topology that we observe here may be quite extreme on account of the unique population distribution in Namibia and could potentially exhibit different patterns in a country with a larger or more evenly distributed population. Future research will therefore benefit from analyzing mobility data from different contexts to establish the best predictors of trip duration that are generalizable and develop a simple data-driven mechanism, which includes these additional sources of heterogeneity, that can be easily incorporated into spatial transmission models.

## Methods

### Geographic data

In addition to the CDR data, we used open source data to provide the static population sizes of each district (*N*_*i*_) and the distances between all districts (*d*_*ij*_). District population sizes were calculated by summing 2010 WorldPop Project (www.worldpop.org) estimates of the total number of people per 100m grid cell within each district (administrative level 2) in Namibia. District-level shapefiles were acquired from DIVA-GIS (www.diva-gis.org) and distances between districts were calculated as the Euclidean distance between district centroids.

### Definition of duration-restricted subnetworks

We performed initial analyses of travel network topology by comparing relative edge weights for travel networks based on three broad intervals of trip duration. Relative edge weights were defined as 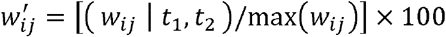 such that each sub-network contained only trips where duration of travel (*t*) was within the interval [*t*_1_, *t*_2_] and is scaled by the overall maximum edge weight observed in the full travel network max(*w*_*ij*_). The broad intervals for this initial analysis were defined as 1–3, 7–14, and 30–60 days in duration (see Figure 2). To further explore how network topology changes on account of trip duration, we subset our data into 20 duration-restricted subnetworks reflecting intervals of trip duration that are analogous to the generation times of several infectious pathogens (e.g. 2-3 days for influenza, 14-21 days for Ebola) with duration intervals continuing up to 12 months (see Table 1).

### Measures of network centrality

To characterize the overall topological structure of each duration-restricted subnetwork in Table 1, we used two centrality measures: node strength (i.e. weighted degree per node) and node closeness (i.e. the cumulative weighted distance separating node *i* from all other nodes)^70^. Here, nodes represent geographic locations (districts). Specifically, we calculated node strength as the out-strength of node *i* as 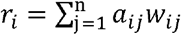, where n is the total number of nodes and *w*_*ij*_ is a matrix of weights given by the total number of trips between *i* → *j* nodes^71^. We calculated node closeness according to Opsahl et al.^70^ using the ‘tnet’ R package^72^, which is defined as 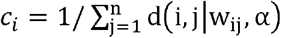, with tuning parameter α= 0.5 so that both edge weight and the number of intermediate nodes are used to calculate the shortest path between two nodes. We then condensed the information from these statistical distributions into two metrics of network heterogeneity.

### Measuring the level of heterogeneity in travel networks

To investigate potential threshold(s) of trip duration at which network topology undergoes significant shifts, we used measures of network heterogeneity and changepoint analysis to evaluate the range of duration-restricted subnetworks for points where there are significant changes in the network structure. Here, a heterogeneous network is characterized by network centrality measures with skewed and/or long-tailed distributions compared to a more homogeneous network which is characterized by centrality measures that have shorter-tailed distributions. A mathematical definition of the level of heterogeneity in a network has been previously described in Barrat et al.^73^ where the first and second moment of a centrality measure’s distribution is concisely translated into a heterogeneity parameter, which we have dubbed η. Following Barrat et al.^73^ we defined the heterogeneity parameter η for the distributions of both node strength and closeness, denoted as *η*_strength_ and *η*_close_ respectively. For example, network heterogeneity as measured by node strength r is:

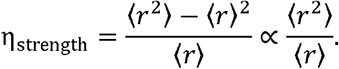

Where, the denominator here represents the expected value of node strength across all *i* nodes 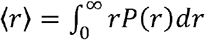 and the numerator represents the variance σ^2^ = ⟨*r*^2^⟩ − ⟨*r*⟩^2^, where the second moment is 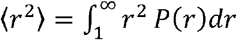. In this case of node strength, we estimated its statistical distribution for all nodes in the given travel network using a kernel density estimator with an equally-weighted mixture of Gaussian kernels^74^ as the probability density function *P*(*r*). Defined as such, η can be considered analogous to the normalized variance of the given centrality measure for the entire travel network. Heterogeneity *η* _strength_ and *η*_close_ are thus heuristic measures that indicate the magnitude of fluctuations in node strength and closeness relative to the mean, such that a highly heterogenous network will have large variance compared to the mean (i.e. *η*_strength_ > ⟨*r*⟩) in comparison with a homogenous one (i.e. *η*_strength_ ≈ ⟨*r*⟩)^73^.

### Estimating shifts in travel network structure

To identify potential thresholds of trip duration at which travel network structure shifts from a heterogeneous to homogenous structure, we performed a multivariate changepoint analysis^75^ based on the joint distribution of the heterogeneity metrics η_*r*_ and η_*c*_ calculated for networks comprised of trips falling within each of the trip duration intervals in Table 1. We estimated the changepoint network heterogeneity using the Energy Divisive algorithm in the ‘ecp’ R package^76^ which estimates both the number and position of changepoints in a multivariate space. The algorithm estimates the *k* changepoints hierarchically by sequentially segmenting observations into groups and iteratively maximizing the divergence of zero-, first-, and second-moment measures among the *k* + 1 groups so that they are mutually independent and identically distributed^77^. We estimated the changepoints by enforcing a minimum group size of 2 with a significance level of 0.05 and 10,000 permutations. Based on the estimated changepoints in (*η*_*r*_, *η*_*c*_) space, we inferred the thresholds of trip duration at which the travel network shifts significantly from heterogeneous to homogeneous.

### Identifying drivers of travel network structure

To explore potential drivers of this shift, we examined the distribution of raw trip counts emanating from each origin and its dependence on two covariates: distance between origin and destination d_*ij*_, and population density of the destination (people per km^2^ calculated as *N*_j_/*A*_*j*_). We fit1log-linear regression models to these trip counts for each origin *i* and trip duration interval [*t*_1_, *t*_2_] in Table 1.

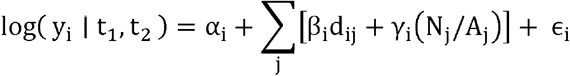

Here, the linear predictor is fitted to the duration-restricted trip counts (*y*_*i*_ | *t*_1_, *t*_2_) using a log link function with origin-level intercept α_i_, coefficients β_i_ and γ_*i*_, and error term ϵ_i_. The models were fit to each origin and duration interval using Maximum Likelihood Estimation. We then compared the effect of the two covariates on duration-restricted travel volume by calculating the effect size (fitted slope) of each origin-specific model and then plotted them against each trip duration interval.

### The impact of trip duration and network structure on spatial disease dynamics

We used a simple model of importation probability to assess the predictability of spatial disease spread under different network structures observed in the travel data from Namibia. We assumed a single introduction of one infected individual in origin location *i* where the proportion of the poplulation susceptible is given by *s*. Exponential growth of infected individuals is given by *I*_*it*_ *=* β ^*t*−1^*s*. The probability of importation from *i* into each of the *j* populations at time *t* is:

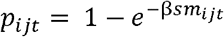

where

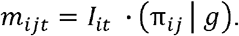

As in most discrete-time models, we make the simplifying assumption that pathogens are infectious for the entirety of the generation time, therefore the transmission rate β is equal to the basic reproductive number *R*_0_. The conditional term (π_ij_ | g) is the fitted value of connectivity π_*ij*_ given the number of observed trips with a duration equal to the generation time *g* of the pathogen (y_ij_ | g). Connectivity was estimated using a gravity model:

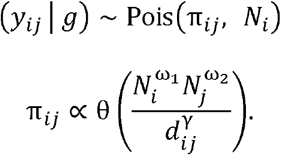

Where, θ is the proportionality constant, ω_1_ and ω_2_ are exponential parameters that control the attractive force between the origin *N*_*i*_ and destination *N*_*j*_ population sizes, and γ scales the distance penalty. Gravity model parameters were estimated using Bayesian inference and Markov Chain Monte Carlo (MCMC) sampling with the ‘mobility’ R package.

To calculate the general distribution of spatial predictability we calculated *p*_*ij*_ for all unique combinations of *R*_0_ from 1 to 30 and generation times from 1 to 30 days. We then calculated the uncertainty in importation to all other potential destinations as the Shannon entropy of the vector *p*_*i*·_ as H(*p*_*i*_) = −∑ *p*_*ij*_log_2_ (*p*_*ij*_).

To more easily compare values of importation uncertainty across all scenarios of *R*_0_ and generation time, we defined the overall predictability of spatial spread from the index location *i* as ϕ_*i*_:

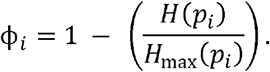

Here the H _max_ (p_i_) function represents the principle of maximum entropy^78^ for a system with an equivalent number of locations n: *p*_*i*·_ = 1/n for all destinations j ∈ {1,…, n}. This formulation gives a value of 0 for the least predictable scenario where probability of importation to all locations is equal (i.e. maximum entropy) and a value of 1 for a perfectly predictable scenario (a single location with importation probability of 1).

## Data Availability

Due to the data-sharing agreement with the mobile phone provider, the authors may not distribute the data used here. These data can be requested directly from MTC mobile (https://www.mtc.com.na/). Code to reproduce analyses is available upon request.

## Declarations

### Ethics approval

The data used in this study was approved by the University of Southampton ERGO committee (ERGO ID 23647). No individuals were enrolled to participate because the data have been aggregated and de-identified.

### Author Contributions

APW, AJT, CJEM, EES collated data. JRG and APW designed the study. JRG performed analyses and wrote first draft of the manuscript. All authors contributed to the final draft of the manuscript.

