## Supplementary Information for "Trip duration drives shift in travel network structure with implications for the predictability of spatial disease spread"

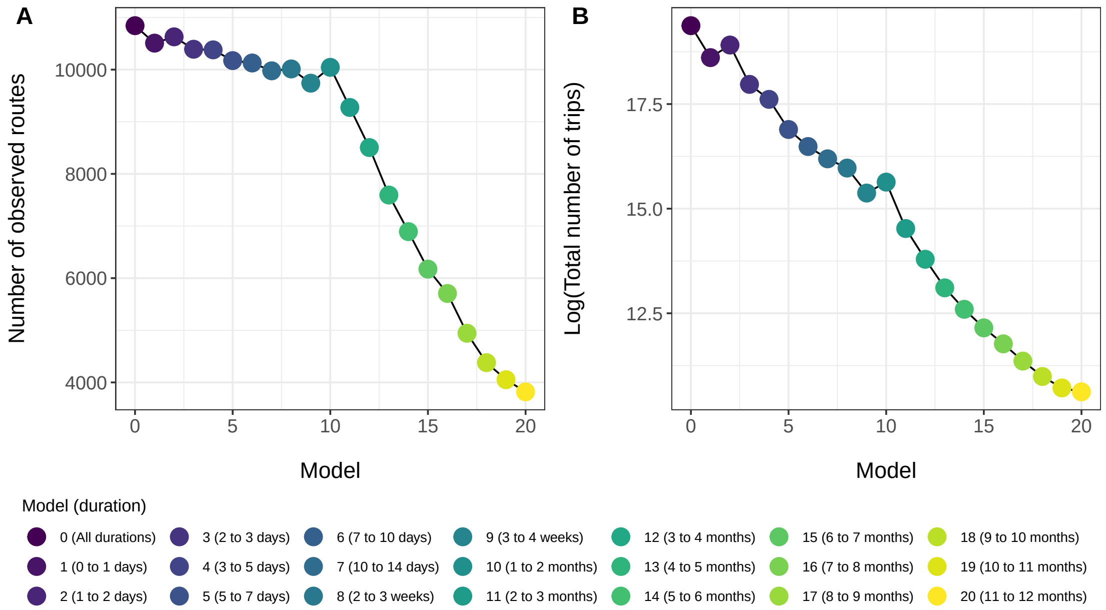


Figure S1: The number of observed routes (A) and the number of observed trips (B) for each of the 20 duration-restricted travel networks.

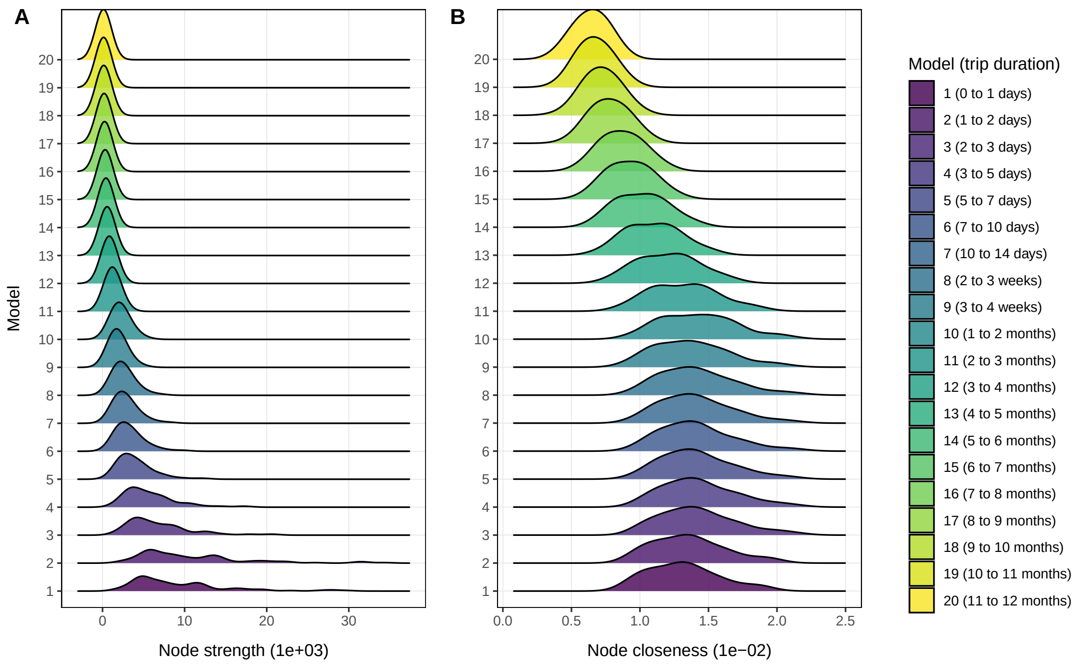


Figure S2: Distributions of node strength (A) and node closeness (B) for each of the 20 duration-restricted travel networks.

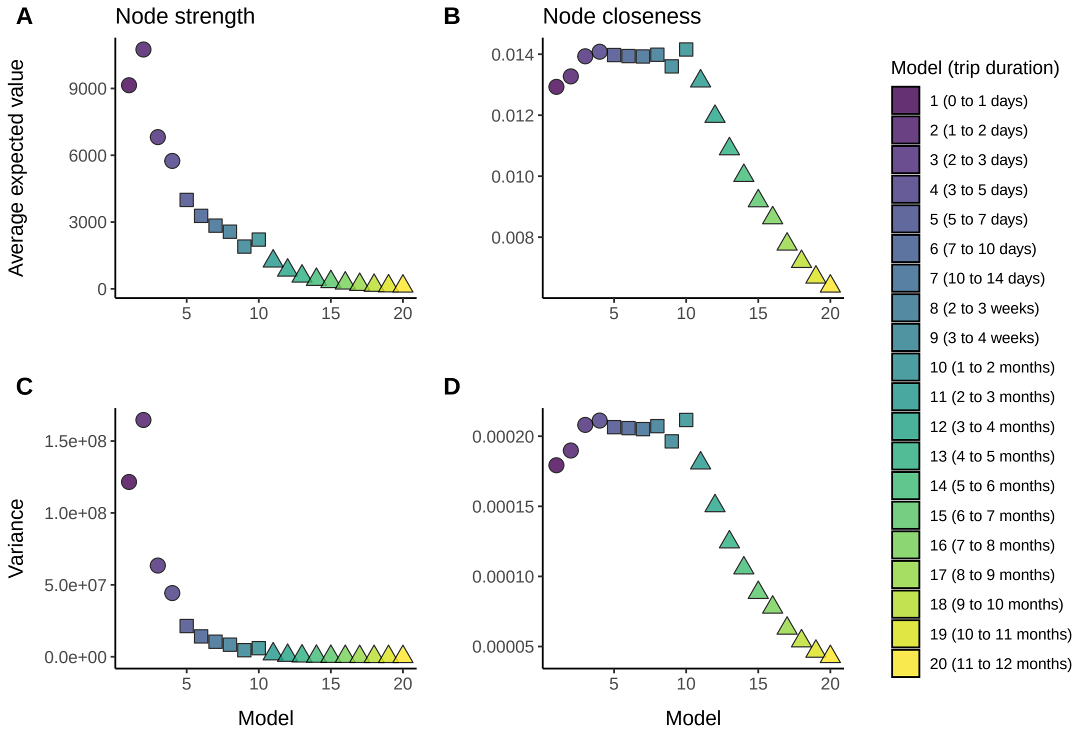


Figure S3: Top: changes in the average expected value of for distributions of node strength $\langle r \rangle$ (A) and node closeness $\langle c \rangle$ (B) across all 20 of the duration-restricted travel networks. Bottom: changes in the variance of distribution of node strength $\langle r^2 \rangle$ (C) and node closeness $\langle c^2 \rangle$ (D) across all 20 of the duration-restricted travel networks.

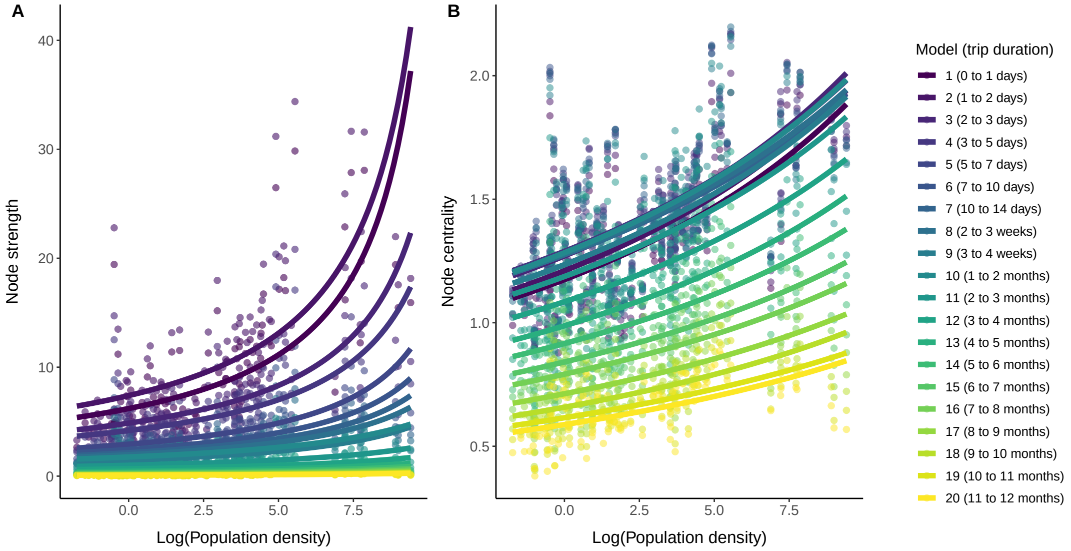


Figure S4: Changes in node strength (A) and node closeness (B) for each duration-restricted travel network according to population density of the origin district. Points show the observed values of node strength and node closeness dependent upon the log population density of each origin district. Trend lines indicate the changes in both networks measures across all values of population density. The color key to the right indicates the duration interval corresponding to each travel network.

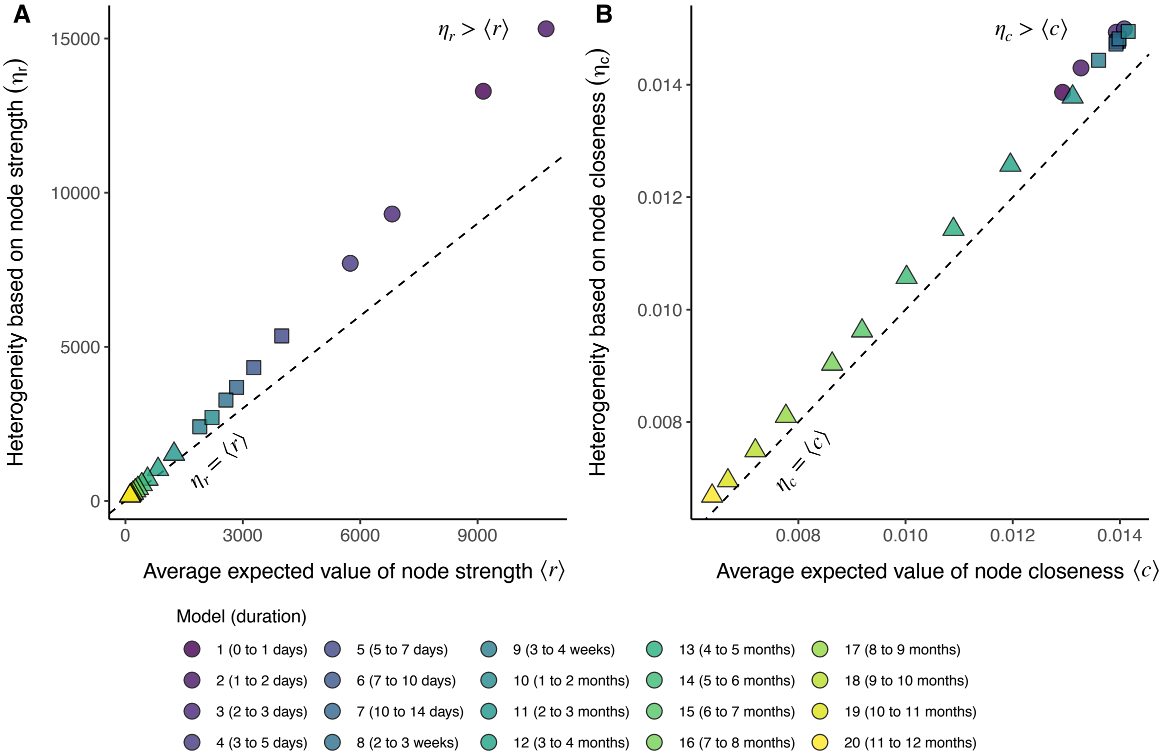


Figure S5: A) The relationship between the network heterogeneity metric based on node strength ($\eta_r$) and the average expected value of the observed distribution of node strength $\langle r \rangle$. B) The relationship between the network heterogeneity metric based on node closeness ($\eta_c$) and the average expected value of the observed distribution of node closeness $\langle c \rangle$. Dashed line indicates the $x = y$ line and each point is colored according to duration-restricted travel network.

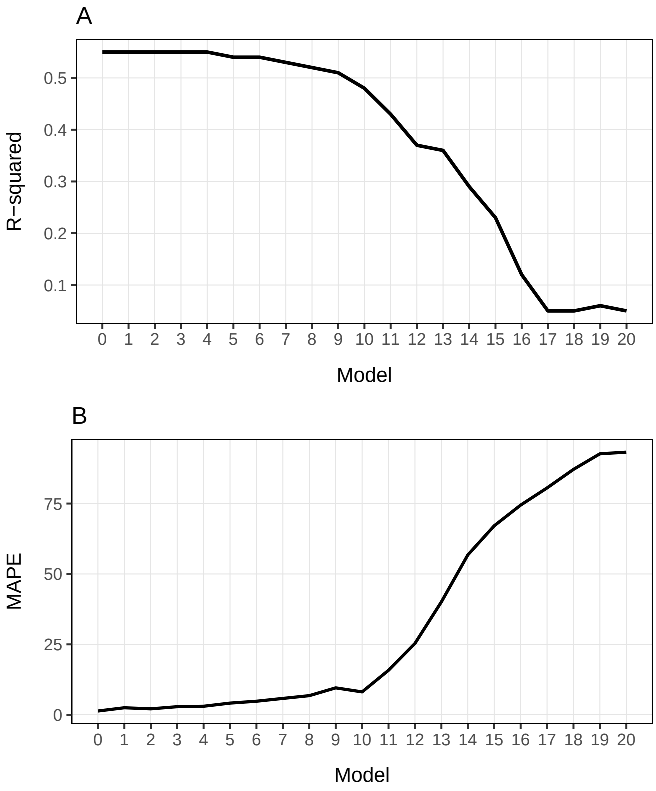


Figure S6: Change in goodness of fit across 20 duration-restrict gravity models. Figure A shows the R-squared value of each model and figure B shows the Mean Absolute Percent Error (MAPE).

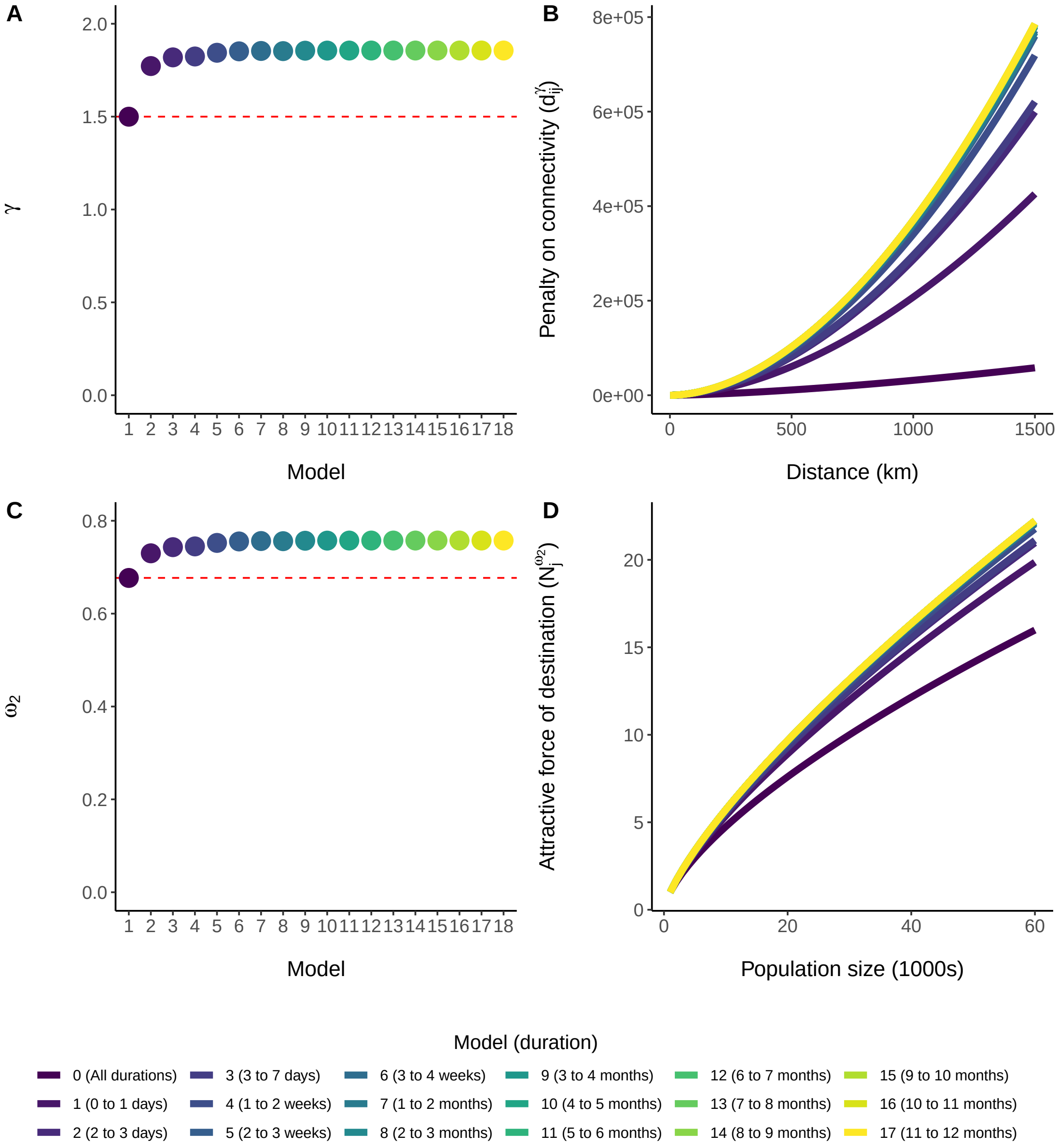


Figure S7: Changes in the gravity model parameters for each randomly down-sampled travel networks. Down-sampled travel networks are the full travel network where the number of observed routes and number of observed trips is sampled randomly according to Table 1 in the main text. Figures A and C show changes in the distance parameter $\gamma$ and population destination size parameter $\omega_2$ respectively. Figure B shows the effective penalty of distance $(d_{ij}^{\gamma})$ on estimated connectivity for each down-sampled network. Figure D shows the change in the attractive force of destination districts $(N_j^{\omega_2})$ based on the population size of the destination for each of the down-sampled networks.

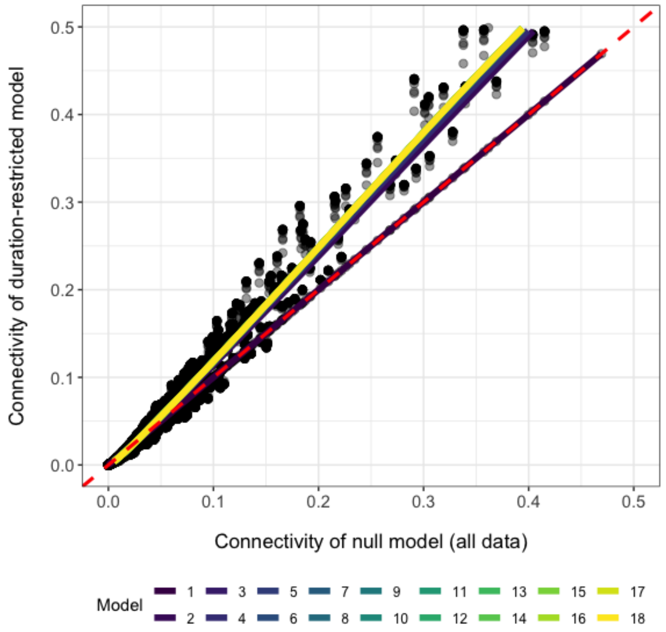


Figure S8: Connectivity values fitted by gravity models for all randomly down-sampled sub-models are compared to connectivity of the full model (all data). Down-sampled travel networks are the full travel network where the number of observed routes and number of observed trips is sampled randomly according to Table 1 in the main text. Red dashed line indicates the $x = y$ relationship.

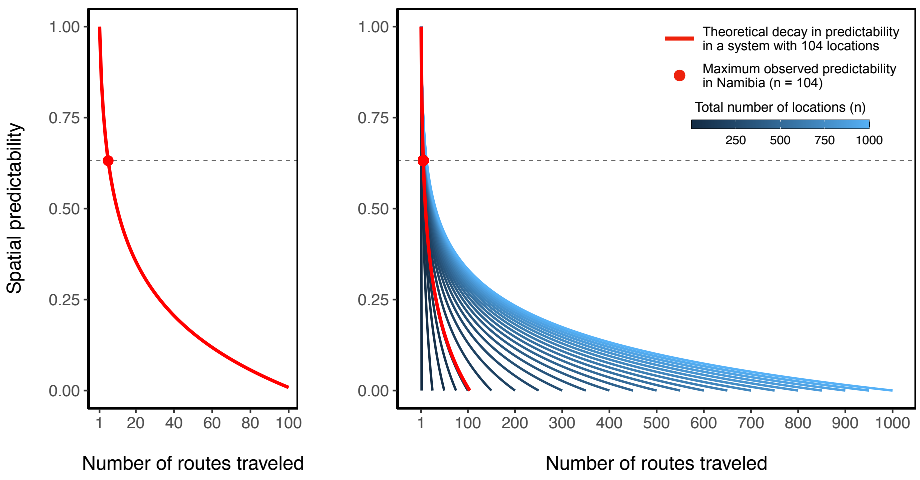


Figure S9: Change in spatial predictability depending on the number of destination locations that are predicted to have disease importation. The left figure shows the theoretical change in spatial predictability for a travel network with one infected origin location and 104 potential destinations ($n = 105$). The red circle indicates the maximum value of spatial predictability ($\phi = 0.63$) based on spatial spread of several pathogens on the Namibia travel network. The figure on the right shows the change in spatial predictability for travel networks of up to $n = 1000$ locations. The travel network size is indicated by the color bar and the network with size equivalent to the Namibia travel network ($n = 105$) is highlighted in red.

Table S1: Transmission parameters for each of the six pathogens used in simulations of spatial spread. Where, $R_0$ is the basic reproduction number, $g$ is the generation time in days, and $s$ is the proportion of the population that is susceptible.

| Pathogen | $R_0$ | $g$ | $s$ | Citation |
|---|---|---|---|---|
| influenza | 2 | 3 | 0.7 | (Anderson and May 1992) |
| SARS-CoV-2 | 3 | 8 | 0.95 | (Anderson et al. 2004) |
| Ebola | 1.5 | 16.6 | 0.95 | (Chowell and Nishiura 2014) |
| measles | 15 | 14 | 0.1 | (Anderson and May 1992) |
| pertussis | 5.5 | 25 | 0.1 | (Beest et al. 2014, Vink et al. 2014) |
| malaria | 10 | 60 | 0.75 | (Huber et al. 2016, Smith et al. 2007) |
